# Adaptive policies for use of physical distancing interventions during the COVID-19 pandemic

**DOI:** 10.1101/2020.05.29.20065714

**Authors:** Reza Yaesoubi, Joshua Havumaki, Melanie H. Chitwood, Nicolas A. Menzies, Gregg Gonsalves, Joshua Salomon, A David Paltiel, Ted Cohen

## Abstract

Policymakers need decision tools to determine when to use physical distancing interventions to maximize the control of COVID-19 while minimizing the economic and social costs of these interventions. We develop a pragmatic decision tool to characterize adaptive policies that combine real-time surveillance data with clear decision rules to guide when to trigger, continue, or stop physical distancing interventions during the current pandemic. In model-based experiments, we find that adaptive policies characterized by our proposed approach prevent more deaths and require a shorter overall duration of physical distancing than alternative physical distancing policies. Our proposed approach can readily be extended to more complex models and interventions.

**One-sentence summaries:** Adaptive physical distancing policies save more lives with fewer weeks of intervention than policies which prespecify the length and timing of interventions.

## Main Text

The health and economic costs of the ongoing SARS-CoV-2 pandemic are staggering. In the absence of an effective vaccine or other pharmaceutical interventions, physical distancing (PD) measures have been the primary means to reduce the speed of epidemic growth, to relieve pressure on the health care system, and to buy scientists time to develop new prevention and treatment strategies. Model-based analysis of previous pandemics (*1*) as well as current models of the COVID-19 pandemic suggest that these PD interventions will need to be maintained for weeks to months in the United States to minimize risks of overwhelming hospital capacity and to avoid epidemic rebound (*2, 3*). However, PD measures also impose an immense economic and social burden. Indeed, it is clear that a single uninterrupted period of strict PD cannot be practically sustained to achieve eradication. Accordingly, policymakers require tools to help them determine when PD interventions should be started and stopped to optimize health and economic outcomes.

Here, we describe a method for adaptive decision making during the ongoing pandemic that explicitly accounts for the tradeoffs between the health benefits and economic costs of PD interventions. We demonstrate how this decision model can assemble the necessary evidence to characterize policies that use surveillance data to inform real-time determination of starting, continuation, or stopping of PD interventions. Using a simulation model of the COVID-19 pandemic, we show that these types of adaptive policies can avert more deaths while requiring a shorter total duration of PD compared to alternative policies, including pre-determined periodic interventions and those that focus on ensuring available ICU capacity is not surpassed (*2, 3*).

For the analysis presented here, we assume that a policymaker’s objectives are to minimize both the number of deaths associated with COVID-19 and the duration of physical distancing interventions as a proxy for the harm inflicted by shutdowns on the economy. We quantify the trade-off between these objectives using the net monetary benefit (NMB) framework (*4, 5*), expressed as 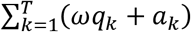, where *T* is the number of weeks the epidemic lasts; *q_k_* is the number of deaths due to COVID-19 that occur in week *k*; and *a_k_* = 1 if physical distancing is in effect during week *k*, and *a_k_* = 0, otherwise.

In the objective function described above, *ω* represents how a specific policymaker weighs the number of COVID-19 deaths per 100,000 population against the duration of PD. For example, a policymaker with trade-off value *ω* = 0.1 weeks per COVID-19 death averted is willing to keep PD in place for one additional week if it could prevent 10 deaths related to COVID-19 per 100,000 population. Higher values of *ω* indicate a higher tolerance for paying the economic and social costs of PD interventions in the interest of reducing COVID-19-related mortality. In the Supplement, we describe how this objective function can be extended to incorporate more comprehensive outcomes including quality-adjusted life-years lost and cost incurred due to cases of COVID-19, and the societal and economic cost of PD in each week.

Here, we estimate *q_k_* using a simple SEIR model of SARS-CoV-2 transmission in a population of size 1,000,000. Informed by data from the COVID-19 pandemic in U.S. (*6*), the model projects the weekly incidence of cases, patients requiring hospitalization and/or critical care, and deaths due to COVID-19. We assume that the PD intervention reduces the population contact rate by 70% with uncertainty interval of [60% − 80%] (*3*). Additional details are provided in the Supplement.

An adaptive policy for informing PD interventions uses surveillance data available at the start of week *k* to recommend whether PD should be started, continued, or stopped during week *k*. One example of such a policy would recommend triggering PD when the estimated prevalence of infection surpasses 375 cases per 100,000 people and stopping PD when this epidemiological measure falls below 100 cases per 100,000 people (*3*). As data such as the weekly number of COVID-19 patients requiring critical care (*C_k_*) may be more easily observed than prevalence of infection, we focus on adaptive policies that use *C_k_* to guide the use of PD interventions during the next week. We consider policies that, for a given trade-off value *ω*, specify two thresholds (*t*_1_ (*ω*), *t*_2_ (*ω*)) which govern the recommendation of PD for the following week. This results in very simple decisions rules: If *C_k_* > *T*_1_(*ω*), PD should be used in the following week and if *C_k_* < *T*_2_(*ω*), PD should be stopped. In the Supplement, we describe the optimization algorithm which identifies the thresholds (*T*_1_(*ω*), *T*_2_(*ω*)) that minimize the loss in the population NMB for a given trade-off value *ω*.

Fig. 1 presents an example of a policy which uses *C_k_* to guide weekly decisions about the use of PD and minimizes the loss in the population NMB. At a trade-off value *ω =* 0.1 weeks per death averted per 100,000 persons, for example, this policy suggests that the PD intervention should be started when *C_k_* exceeds a threshold 13.2 per 100,000 persons (Fig. 1A) and stopped when *C_k_* falls below 12.9 per 100,000 persons (Fig. 1B). For a policymaker with an uncertain trade-off value *ω*, Fig. 1 presents an affordability curve (Fig. 1C), which returns the cumulative number of weeks for which PD interventions are expected to be used for different values of *ω*. A policymaker with a maximum tolerance for the overall number of weeks of PD can consult Fig. 1C to select a trade-off threshold that satisfies this constraint in expectation. The policymaker can then use Fig. 1A-B to make real-time decisions given their selected trade-off threshold. We note that higher trade-off values lead to lower thresholds of *C_k_* at which to start PD interventions (Fig. 1A-B), more weeks during which PD is used (Fig. 1C) and a lower expected number of deaths due to COVID-19 over the course of the pandemic (Fig. 1D).

**Fig. 1:**
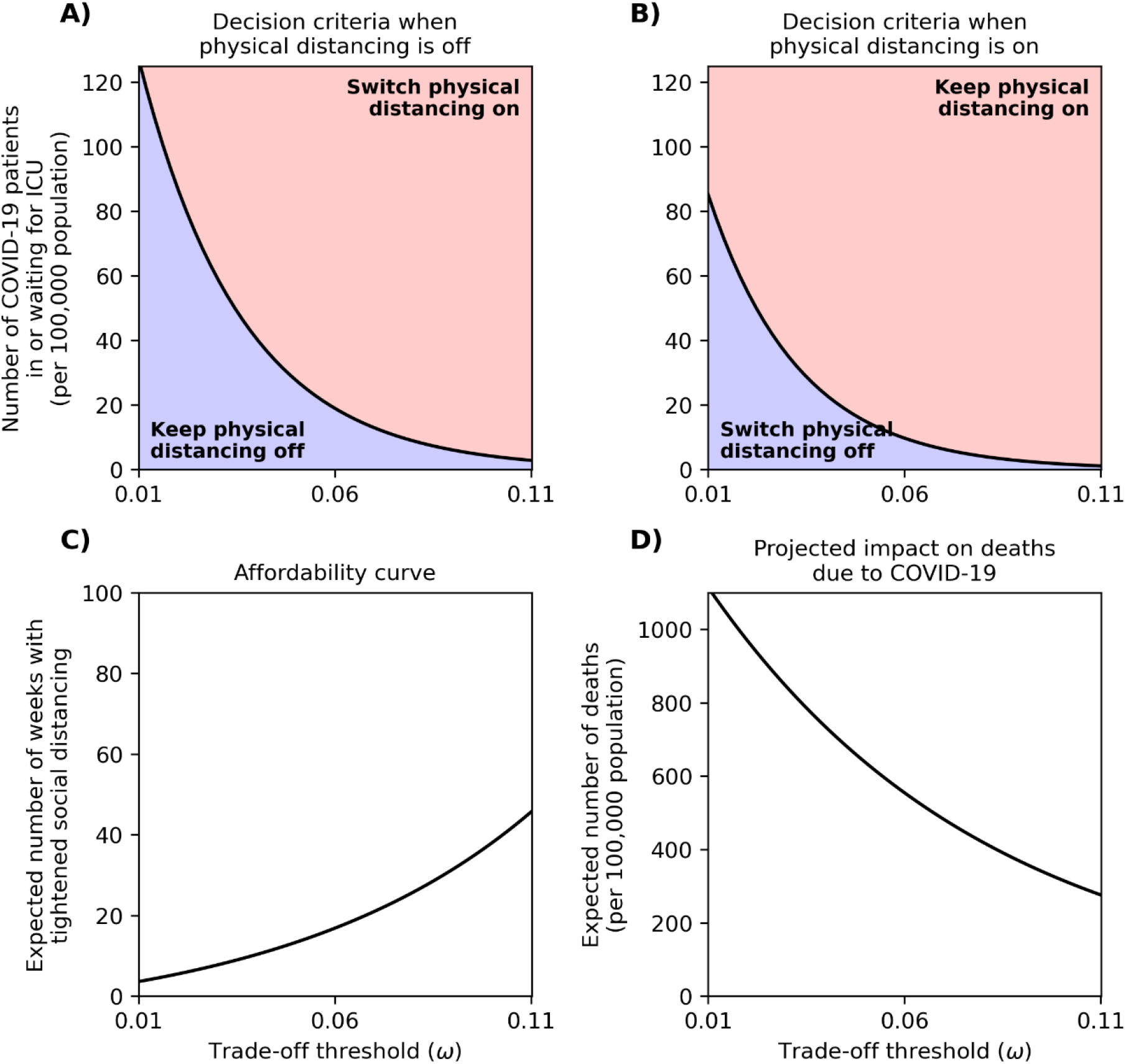
A policy that uses the number of COVID-19 patients who need critical care (*C_k_*) at the beginning of a given week to inform whether the physical distancing interventions should be turned on or off during the following week. Panel A displays the decision rule to be used when the physical distancing intervention is currently off and panel B displays the decision rule when the physical distancing intervention is currently on. Panel C returns the accumulated number of weeks during which physical distancing is expected to be used, and Panel D displays the number of deaths per 100,000 population expected to occur due to COVID-19 under adaptive policies that correspond with different trade-off values (*ω*) (Panels A-B).

As we have shown in previous studies of epidemics (*7-9*), the performance of policies to guide decision-making depends on the specific features of surveillance data (e.g. *C_k_*) selected to inform decisions. To demonstrate the differential performance of PD policies in the context of the ongoing epidemic, we used our simulation model of the COVID-19 pandemic to simulate the outcomes of three types of PD policies that use different features and decision rules to inform recommendations (Fig. 2). We consider a *static* periodic policy (purple curve) that employs the periodic use of PD at pre-defined durations and frequencies (e.g. every 2 weeks for 2 weeks, every 4 weeks for 4 weeks, etc.). We also evaluate two *adaptive* policies that use *C_k_* to determine whether PD should be started, continued or stopped for the following week: 1) an ‘Adaptive: ICU Capacity’ policy which is similar to policies proposed by (*2, 3*) where the on/off PD thresholds are determined to ensure that the expected probability of surpassing available ICU capacity is below a certain value (e.g. 90%); and 2) an ‘Adaptive: Minimize Loss of NMB’ that determines the on/off thresholds to minimize the loss in the population NMB using the optimization algorithm described in the Supplement (this policy is displayed in Fig. 1A-B).

**Fig. 2:**
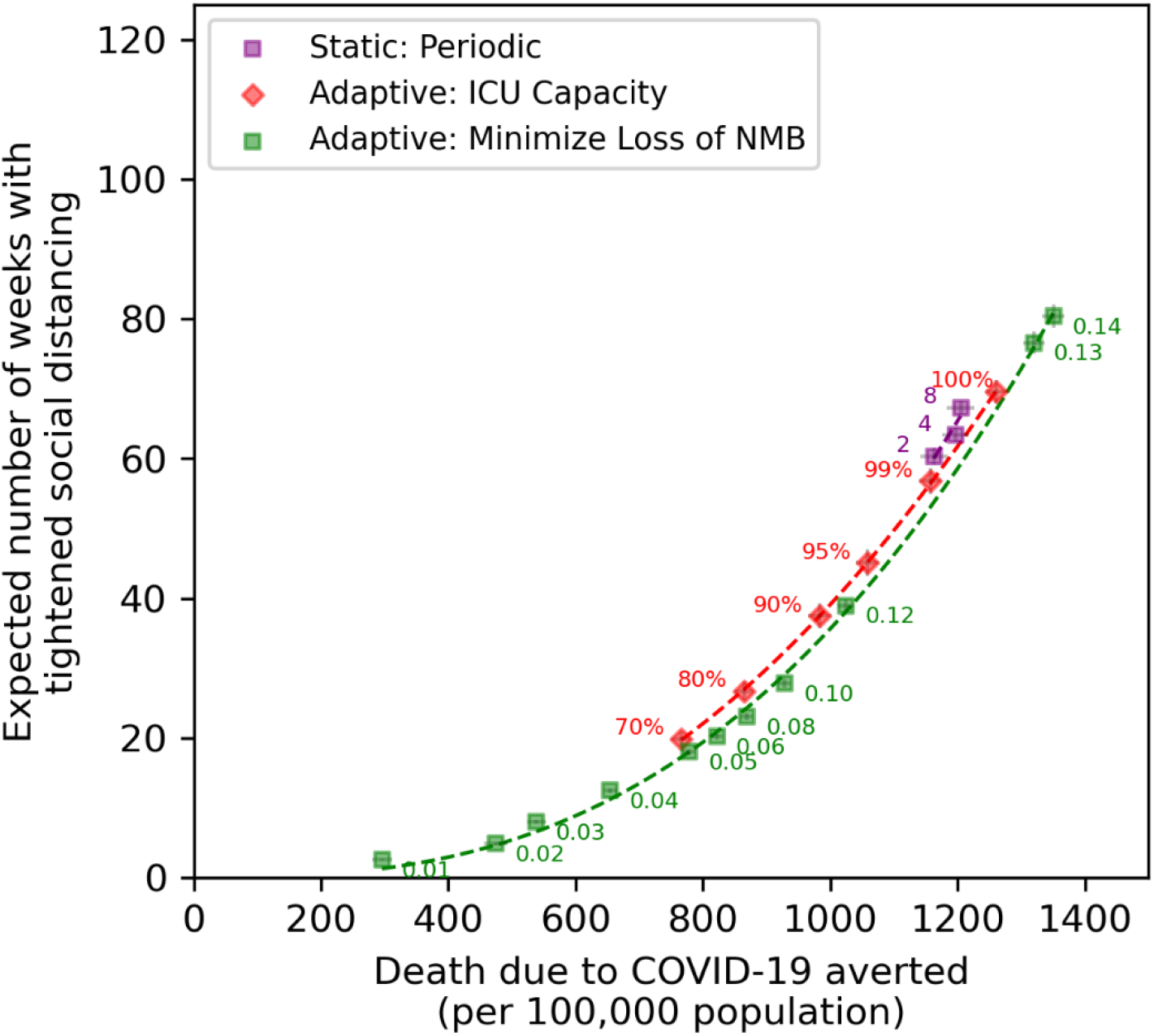
Comparing the performance of difference physical distancing (PD) policies. The origin in this figure corresponds to a counterfactual scenario where PD is not used at all, which is expected to result in 1,391 deaths per 100,000 population. The curve ‘Static: Periodic’ corresponds to policies that recommend using PD every 2 weeks for 2 weeks, every 4 weeks for 4 weeks, and so on. The numbers on the red curve (‘Adaptive: ICU Capacity’) represent the probability that a COVID-19 patient who requires critical care would get admitted to ICU. The numbers on the green curve (‘Adaptive: Minimize Loss of NMB’) represent the trade-off thresholds of policies selected from Fig. 1A-B. The bars represent the 95% confidence interval of projections using 200 simulated trajectories. NMB: net monetary benefit.

The adaptive policies that seek to minimize the loss in NMB can be designed to *dominate* (i.e. prevent more deaths and require shorter duration of PD as) other policies described above (Fig. 2). For any given number of weeks the policy maker is willing to maintain the PD intervention, the horizonal distance between the green curve and other curves in Fig. 2 represents how many deaths could be prevented by using an adaptive policy designed to minimize the loss in NMB, as compared to one of the other two approaches.

The results presented here highlight the need for several specific data items to promote better decision-making during this pandemic. First, we need the best data possible to understand the natural history of the pathogen (e.g. duration of immunity, infectiousness, role of asymptomatic transmission, mortality), the current epidemiology (e.g. the prevalence of infectious individuals and hospitalized patients), and the effectiveness (e.g. reduction in contact rate) of PD interventions. Second, we need better data on the economic costs of physical distancing interventions. Third, and most difficult, is the need for policymakers to establish what economic tradeoffs we are willing to make to preserve the health of the population. In the absence of clearly established guidelines on societal willingness-to-pay for health, policymakers may choose some proxies for the consumption of societal resources such as the number of weeks over which PD interventions are expected to be used (Fig. 1C) to choose an efficient PD policy to follow.

This paper presents a simple model of the COVID-19 pandemic in order to facilitate exposition and discussion. As such, we have made a number of assumptions. We average across many important sources of heterogeneity. Additional details (e.g. age-structure, risk-group membership, asymptomatic transmission, population compliance with different PD interventions over time, availability of tests, and lags in the reporting of test results) will need to be added to allow for context-specific usage and to allow for simulation of more complex interventions (e.g. contact tracing, age-specific relaxation of PD rules, use of serological tests to establish which individuals should not be limited by PD interventions). We used crude estimates to project health loss associated with the COVID-19 pandemic ignoring comorbidities and downstream health consequences of unemployment during tightened PD interventions. These factors could also be incorporated, depending on the availability of data, in the objective function we described above and in the Supplement. The effectiveness of different PD interventions may not be known a priori, and our simple approach for modeling PD ignores the complexities in which mobility may change over the course of the epidemic regardless of the PD policy adopted. New data on the relationship between mobility, contact patterns, and the types of PD interventions guidelines in place can help improve the usefulness of these models.

The decision tool we present here provides a flexible framework for using real-time observations to guide the use of PD interventions. It seems likely to us that policymakers would prefer policies that are responsive to updated surveillance and do not prematurely commit them to future actions. The algorithm we propose to optimize PD policies does not restrict the type of epidemic model used to project the health and economic outcomes of the COVID-19 pandemic. Various frameworks, including compartmental, agent-based, or network models may be used as needed to relax the simplifying assumptions of our model. Furthermore, our proposed approach can also incorporate delays in reporting of data and uncertainties in estimates (due to limited sample size) (*7, 9*). We have described an algorithm to optimize PD policies in the Supplement and welcome other groups to contact us or adapt this approach to help improve real-time decision making during this epidemic.

## Data Availability

All data needed to replicate this study is included in the main text and the supplement.

## Acknowledgments

RY was supported by K01AI119603, and TC by R01AI112438 and R01AI146555, all from the National Institute of Allergy and Infectious Diseases. GG was supported by DP2DA49282 and ADP by R37DA015612 both from the National Institute on Drug Abuse. Author contributions: RY: Conceptualization, Formal Analysis, Investigation, Methodology, Software, Visualization, Writing (original draft); JH: Data curation, Writing (review and editing); MC: Data curation, Writing (review and editing); NAM: Supervision, Writing (review and editing); GG: Supervision, Writing (review and editing); JS: Supervision, Writing (review and editing); ADP: Supervision, Writing (review and editing); TC: Conceptualization, Funding acquisition, Methodology, Supervision, Writing (original draft). Authors declare no competing interests. All data is available in the main text or the supplementary materials. The code of the model is provided at https://github.com/yaesoubilab/APACE.

**Supplementary Materials:** Materials and Methods, Figures S1, and Tables S1-S2.

## Notes

### Competing Interest Statement

The authors have declared no competing interest.

